# Causal mediation for uncausally related mediators in the context of survival analysis

**DOI:** 10.1101/2024.02.16.24302923

**Authors:** Arce Domingo-Relloso, Allan Jerolon, Maria Tellez-Plaza, Jose D. Bermudez

**Author notes:** **Corresponding author:** Arce Domingo-Relloso, PhD, Department of Biostatistics, Columbia University Mailman School of Public Health, 722 West 168th Street, 10032 New York, NY. Equal author contribution (Maria Tellez-Plaza, Jose D. Bermudez).

## Abstract

**Objective:** The study of the potential intermediate effect of several variables on the association between an exposure and a time-to-event outcome is a question of interest in epidemiologic research. However, to our knowledge, no tools have been developed for the evaluation of multiple correlated mediators in a survival setting.

**Methods:** In this work, we extended the multimediate algorithm, which conducts mediation analysis in the context of multiple uncausally correlated mediators, to a time-to-event setting using the semiparametric additive hazards model. We theoretically demonstrated that, under certain assumptions, indirect, direct and total effects can be calculated using the counterfactual framework with collapsible survival models. We also adapted the algorithm to accommodate exposure-mediator interactions.

**Results and conclusions:** Using simulations, we demonstrated that our algorithm performs better than the product of coefficients method, even for uncorrelated mediators. The additive hazards model quantifies the effects as rate differences, which constitute a measure of impact, with applications that can be highly informative for public health. Our algorithm can be found in the R package *multimediate*, which is available in Github.

## 1 Introduction

The understanding of causal pathways underlying the association between an exposure or treatment and an outcome is a question of interest in epidemiologic research. Mediation analysis aims to quantify to which extent the relationship between two variables happens through a third variable called the mediator (indirect effect), and to which extent it happens through other not considered pathways (direct effect). Extensive literature, as well as many analytic tools, exist for the evaluation of simple mediation analysis [1, 2].

Nevertheless, the fact that the effect of an exposure or treatment on an outcome will happen through only one mediating variable is unlikely in practice. Some work has been conducted for settings in which multiple mediators exist [3, 4, 5, 6]. The identification of the joint indirect effect for all mediators is straightforward, however, individual indirect effects cannot be identified when conducting individual mediation models for each mediator in the settings in which mediators are correlated. Jerolon et [7] recently developed a quasi-bayesian algorithm to conduct multiple mediation analysis in the setting of uncausally correlated mediators. They implemented this algorithm in the R package *multimediate* for linear and binary outcomes.

On the other hand, interactions between the exposure and the mediator on the association with the outcome are common in practical settings [8]. One of the main advantages of the counterfactual mediation framework [9] as compared to traditional mediation methods is that it can accommodate interactions between the exposure and the mediator.

Moreover, the study of the effect of exposure or treatment variables on time-to-event outcomes (i.e. survival outcomes) is a common research question in epidemiology. Cox proportional hazards models are the most widely used in epidemiologic research. However, due to the lack of collapsibility of the hazard ratio [10, 11], these models are not, in general conditions, the most suitable for mediation analysis. The *mediation* R package, the most widely used statistical package to conduct mediation analysis, uses accelerated failure time models, which are, similarly, non-collapsible [12]. Additive hazards models [13] have also been used to conduct mediation analysis in a survival setting. These models quantify the effects on an additive scale (as rate differences), thus providing a more interpretable measure of impact that can be highly informative for public health [14].

In this work, we extended the method proposed by Jerolon et al. [7] to the survival analysis setting using additive hazards models. We extended the code of the *multimediate* R package, and provide the generalization to survival analysis of the theoretical results previously proved in Jerolon et al. [7] for continuous and binary outcomes. We additionally adapted the multimediate algorithm to accommodate exposure-mediator interactions. We prove the performance of the algorithm for survival analysis using a simulation study comparing the results obtained using the multimediator algorithm to those obtained using simple mediation with the product of coefficients method. The multimediate function for survival has been included in the Github repository *https://github.com/AllanJe/multimediate*.

## 2 Background and notation

Mediation analysis aims to disentangle the extent to which an association between an exposure or treatment and an outcome is partly or totally attributable to a third intermediate random variable, called the mediator. We denote *E* as an exposure or treatment, *M* as the mediator, which is dependent on the exposure *E*; *X* as a set of covariates and *Y* as the outcome of interest. Following the counterfactual framework [15], we consider *Y* (*e*^*∗*^, *M* (*e*)) as the counterfactual outcome, i.e., the value the outcome would take had the exposure been set to *e*^*∗*^ and the mediator been set to the value it would take when the exposure is set to *e*. Following [16, 7, 17], we define the average indirect effect of changing the exposure from *e*^*∗*^ to *e* when the covariates are set to *X* = *x* as follows:

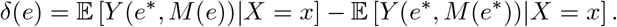

Similarly, the average direct effect, which refers to the effect of the exposure or treatment on the outcome which does not happen through the mediator, is quantified as:

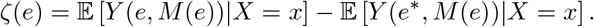

Please note that there is an indirect and a direct effect for each *e*. Last, the average total effect, which denotes the effect of the exposure or treatment on the outcome both through the mediator pathway and through other pathways, is quantified as:

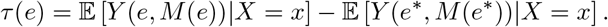

Please note that, following these definitions, it holds that *τ* (*e*) = *ζ*(*e*) + *δ*(*e*), showing that the indirect and direct effects represent an exact decomposition of the total effect. The Sequential Ignorability Assumptions, related to no unmeasured confounding in the exposure-outcome, exposure-mediator and mediator-outcome relationship, in addition to the assumptions of positivity and consistency, need to hold for these effects to be identified [18].

### 2.1 Simple mediation analysis in survival settings

#### 2.1.1 Additive risks model

Although the Cox proportional hazards model is the most widely used in survival analysis, the coefficients of this model represent the log hazard-ratio, which is typically exponentiated to obtain the hazard ratio; a measure that represents the risk of having an event in time *t* given that the event did not happen before. The hazard ratio is a non-collapsible measure [19], which implies that conditioning on a covariate that is related to the outcome would change the coefficient of the exposure, even if the covariate is unrelated to the exposure. Therefore, the hazard ratio of an exposure with the mediator in the model versus without the mediator in the model are not directly comparable, and traditional mediation methods such as the difference of coefficients or product of coefficients methods [20] cannot be used to validly estimate direct and indirect effects. Conversely, measures from additive models are collapsible. For this reason, additive hazards models have been widely used in mediation analysis instead of Cox proportional hazards models or accelerated failure time models.

The Aalen additive hazards model [21] assumes that the hazard function (or the rate) for the failure time *t*, dependent on an exposure *E*, a mediator *M* and a covariates matrix *X*, takes the form:

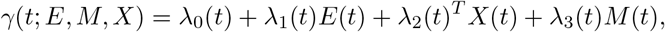

being *λ*_0_ the baseline hazard. Lin and Ying [22] developed the semi-parametric additive risks model, in which the same form of the hazard function is assumed, but the covariates and coefficients can have either time-varying or constant effects. For this work, we will focus on time-invariant covariates and coefficients for simplicity, therefore using the Lin-Ying model, or additive risks model. Only the baseline hazard *λ*_0_ is dependent on time, and the hazard function would then be:

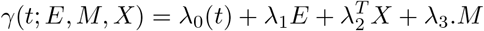

#### 2.1.2 Effect definition

Lange and Hansen [17] defined the direct, indirect and total effects in a survival context for a single mediator considering an additive hazards model as the outcome model, in which the rate *γ* is taken as the outcome. Let us assume that the mediator is continuous and use a linear model for the mediator model. Thus, being *E* the exposure, *M* the mediator and *X* a vector of *p* covariates, the outcome and mediator models are defined as follows:

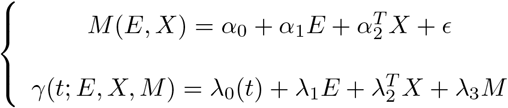

where *α*_0_, *α*_1_, *λ*_1_, *λ*_3_ *∈* ℝ ; *α*_2_, *λ*_2_ *∈* ℝ ^*p*^; *λ*_0_(*t*) is the time-varying baseline hazard and *ϵ* ∼ *𝒩* (0, *σ*^2^) is the error in the linear model, with variance *σ*^2^. The first equation is called the mediator model, whereas the second one is referred to as the outcome model.

In survival settings, the mediated effect, or indirect effect of changing the exposure from *e*^*∗*^ to *e*, is quantified as:

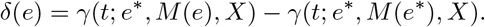

The direct effect is quantified as:

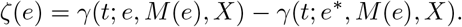

Last, the total effect is quantified as:

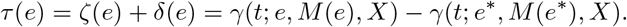

Please note that, here, the effects are defined as differences in hazard functions instead of differences of averages. In general, *δ*(*e*), *ζ*(*e*) and *τ* (*e*) are functions of *t*.

#### 2.1.3 Sequential Ignorability Assumptions in survival analysis

We define *T* (*e, m*) as the time to event when the exposure is set to *e* and the mediator is set to *m*. The following assumptions are sufficient conditions for the direct, indirect and total effects to be identifiable:

- H.1.1. First exchangeability assumption: No unmeasured confounding of the exposure-outcome relationship: *E ⊥ T* (*e, m*) | *X*.
- H.2.1. Second exchangeability assumption: No unmeasured confounding of the mediator-outcome relationship : *M ⊥ T* (*e, m*) | *X, E*.
- H.3.1. Third exchangeability assumption: No unmeasured confounding of the exposure-mediator relationship: *E ⊥ M* (*e*) | *X*.
- H.4.1. Consistency: *M* (*e*) = *M, T* (*e, m*) = *T*.
- H.5.1. *M* (*e*^*∗*^) *⊥ T* (*e, m*) | *X*.

In *Theorem 1* of Lange and Hansen [17], it was proven that, under sequential ignorability assumptions, the total effect measured in the rate difference scale at time *t* is:

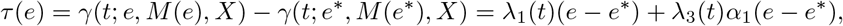

where *λ*_1_(*t*)(*e−e*^*∗*^) is the direct effect of the exposure on the outcome, and *λ*_3_(*t*)*α*_1_(*e−e*^*∗*^) is the indirect efect through the mediator. The proof of this result can be found in the Appendix of Lange and Hansen [17]. Please note that, if *λ*_1_ and *λ*_3_ are time-independent, the three effects wil also be time-independent.

### 2.2 Multiple mediation analysis

Imai and Yamamoto [5] extended the effect definition for simple mediation analysis to the multiple mediators setting. Let us assume that *Z* = (*M*_1_, …, *M*_*K*_)^*T*^ is the vector of all mediators, with *K ≥* 2. Considering *M*_*k*_ as the mediator of interest, *k* = 1, …, *K*, let us define *W*_*k*_ as the vector of all mediators except *M*_*k*_. We also consider *Y* (*e*^*∗*^, *M*_*k*_(*e*), *W*_*k*_(*e*^*∗*^)) as the counterfactual outcome, i.e., the value the outcome would take had the exposure been set to *e*^*∗*^, the mediator of interest been set to the value it would take when the exposure is set to *e* and the other mediators been set to the value they would take when the exposure is set to *e*^*∗*^. In the multiple mediator setting, with *K ≥* 2 mediators, the average mediated effect of the *k*-th mediator is given by:

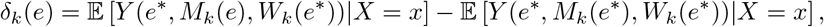

being *X* the covariate vector. The joint indirect effect of all mediators is defined as:

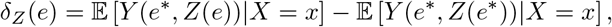

being *Z* the vector of all mediators. The direct effect is defined as:

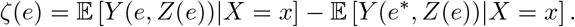

Last, the total effect is defined as:

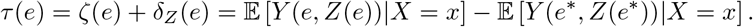

Jerolon et al. [7] defined the direct and indirect effects for continuous and binary outcomes in multiple mediation settings with uncausally correlated mediators. As in simple mediation analysis, in order for the direct, indirect and total effects to be identifiable in multiple mediators settings, several assumptions need to hold. The authors rely on the following hypothesis.

#### 2.2.1 Sequential Ignorability for Multiple Mediators Assumptions (SIMMA)

We define *Y* (*e, m, w*) as the value the outcome would take when the exposure is set to *e* and the mediator is set to *m*.

- H.1.2. *{Y* (*e, m, w*), *M* (*e*^*∗*^), *W* (*e*^*∗∗*^)*} ⊥ E*|*X* = *x*.
- H.2.2. *Y* (*e*^*∗*^, *m, w*) *⊥* (*M* (*e*), *W* (*e*))|*E* = *e, X* = *x*
- H.3.2. *Y* (*e, m, w*) *⊥* (*M* (*e*^*∗*^), *W* (*e*))|*E* = *e, X* = *x*

In addition, the authors assume both the positivity assumption: *P* (*E* = *e*|*X* = *x*) *>* 0 and *P* (*M* = *m, W* = *w*|*E* = *e, X* = *x*) *>* 0 *∀x, e, e*^*∗*^, *m, w*; and the Stable Unit Treatment Value Assumption (SUTVA), which implies that:

1. Potential mediator and outcome values of individual *i* are not dependent on exposures of other individuals, i.e.: *M*_*ik*_(*E*) = *M*_*ik*_(*E*_*i*_) and *Y*_*i*_(*E, M*_*k*_, *W*_*k*_) = *Y*_*i*_(*E*_*i*_, *M*_*ik*_, *W*_*ik*_).
2. There are no multiple versions of exposures, i.e. 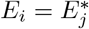 implies 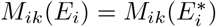 and 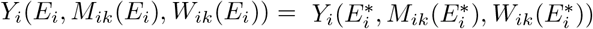
3. There are no multiple versions of mediators, i.e. if 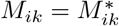, then 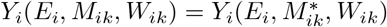.

#### 2.2.2 Multiple mediation analysis for continuous outcomes

In the case of continuous outcomes and *K* independent or uncausally correlated mediators, Jerolon et al. [7] assume the following linear models for both the mediators and the outcome:

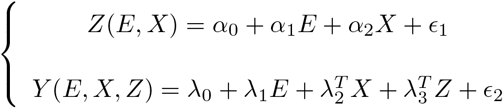

where *α*_0_, *α*_1_, *λ*_3_ *∈* ℝ^*K*^, *α*_2_ *∈* ℝ^*K*^ *×* ℝ^*p*^, *λ*_2_ *∈* ℝ^*p*^, *λ*_0_, *λ*_1_ *∈* ℝ, *ϵ*_1_ ∼ *𝒩*_*K*_(0, Σ) is the vector of residuals with covariance matrix Σ *∈* ℝ^*K*^ *×* ℝ^*K*^, and *ϵ*_2_ ∼ *𝒩*_*K*_(0, *σ*^2^), with *σ*^2^ *∈* ℝ.

Under SIMMA, *Corolary 3.2* in Jerolon et al. [7] shows that the indirect effect of the *k*-th mediator is given by:

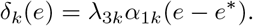

In addition, the joint indirect effect of all mediators is given by:

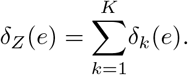

Last, the direct effect is given by:

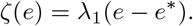

#### 2.2.3 Multiple mediation analysis for binary outcomes

In the case of binary outcomes and *K* independent or uncausally correlated mediators, Jerolon et al. [7] assume linear models for the mediators and a logistic or probit model for the outcome. Assuming a logistic regression model for the outcome:

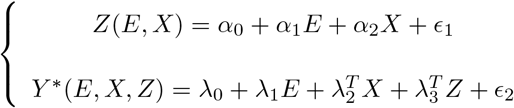

where 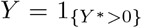, *α*_0_, *α*_1_, *λ*_3_ *∈* ℝ^*K*^, *α*_2_ *∈* ℝ^*K*^ *×* ℝ^*p*^, *λ*_2_ *∈* ℝ^*p*^, *λ*_0_, *λ*_1_ *∈* ℝ, *ϵ*_1_ ∼ *𝒩*_*K*_(0, Σ) is the vector of residuals with covariance matrix Σ *∈* ℝ^*K*^ *×* ℝ^*K*^, and *ϵ*_2_ ∼ *𝒩*_*K*_(0, *σ*^2^), with *σ*^2^ *∈* ℝ.

Under SIMMA, *Corolary 3.3* in Jerolon et al. [7] shows that the indirect effect of the *k*-th mediator is given by:

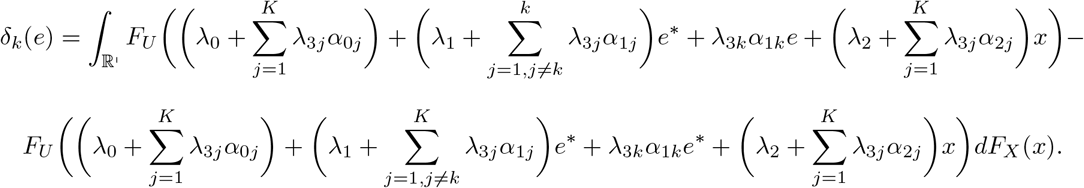

In addition, the joint indirect effect of all mediators is given by:

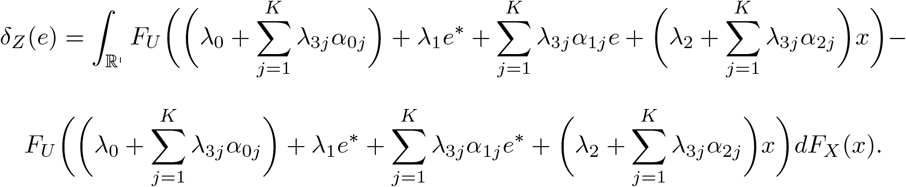

Last, the direct effect is given by:

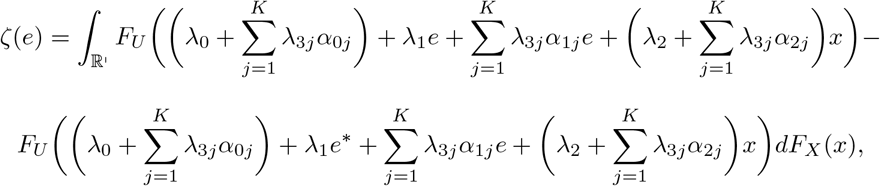

where

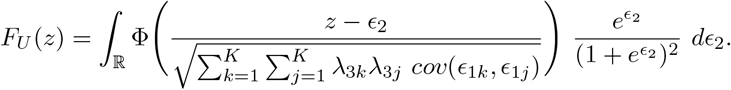

The proof of these expressions can be found in Jerolon et al. [7].

In the following section, we extend these results to the case of survival outcomes.

## 3 Multiple mediation in survival analysis

### 3.1 Effect definition

Following Lange and Hansen 2011 [17] and Jerolon et al. 2021 [7], we define the indirect effect of the mediator *M*_*k*_, *k* = 1, …, *K* as:

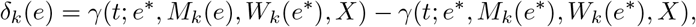

being *γ* the hazard, or rate, function, which is given, for each (*e, e*^*∗*^, *e*^*∗∗*^), by:

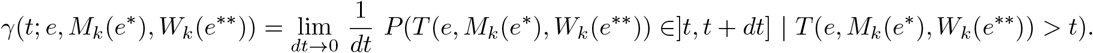

We define the joint indirect effect of all mediators as:

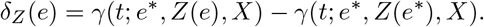

The direct effect is defined as:

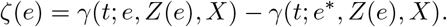

Last, the total effect is defined as:

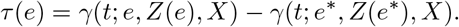

By the above definitions, *τ* (*e*) = *δ*_*Z*_(*e*) + *ζ*(*e*).

### 3.2 Hypothesis

Adapting Lange and Hansen (2011)’s hypothesis [17] to Jerolon et al.’s notation [7], the following set of assumptions is obtained. Let us consider *T* (*e, m, w*) as the time to event when the exposure is set to *e*, the mediator of interest is set to *m* and the other mediators are set to *w*.

- H.1.3: *E ⊥* (*T* (*e, m, w*), *M*_*k*_(*e*^*∗*^), *W*_*k*_(*e*^*∗∗*^)) | *X, ∀k* = 1, …, *K*.
- H.2.3: *T* (*e*^*∗*^, *m, w*) *⊥* (*Z*(*e*)) | *X, E*.
- H.3.3: *T* (*e, m, w*) *⊥* (*M*_*k*_(*e*^*∗*^), *W*_*k*_(*e*^*∗∗*^)) | *X, E, ∀k* = 1, …, *K*.
- H.4.3: *M*_*k*_(*E*) = *M*_*k*_, *W*_*k*_(*E*) = *W*_*k*_, *T* (*E, Z*) = *T*.

We also assume that *P* (*E* = *e*|*X* = *x*) *>* 0 and *P* (*M* = *m, W* = *w*|*E* = *e, X* = *x*) *>* 0 *∀ e, e*^*∗*^, *x, m, w*; and that SUTVA holds.

In addition to SIMMA and SUTVA, we assume that the mediators follow a multivariate multiple linear normal homoscedastic model, and that hazard functions follow the additive risks model, with time-independent coefficients. Therefore, the outcome and mediator models in survival settings with multiple mediators are defined as follows:

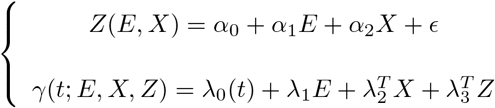

where *α*_0_, *α*_1_, *λ*_3_ *∈* ℝ^*K*^, *α*_2_ *∈* ℝ^*K*^ *×* ℝ^*p*^, *λ*_2_ *∈* ℝ^*p*^, *λ*_1_ *∈* ℝ, *λ*_0_(*t*) is the time-varying baseline hazard and *ϵ* ∼ *𝒩*_*K*_(0, Σ) is the error vector of the multivariate linear regression, with covariance matrix Σ *∈* ℝ^*K*^ *×* ℝ^*K*^.

We also assume, following Jerolon et al. 2020 [7], that, either the mediators are independent, or the correlations between the *k* mediators are not causal, i.e., that the dependence between them does not have a causal order. In this latter case, we assume that pairwise correlations between mediators are independent of the exposure:

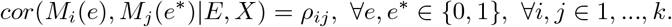

#### Proposition 1

*Under the previous conditions, it holds that the hazard function takes the following value:*

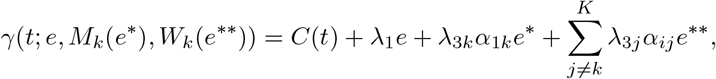

*∀*(*e, e*^*∗*^, *e*^*∗∗*^) *∈ {*0, 1*}*^3^, *being C*(*t*) *a function that does not depend on the exposure values e, e*^*∗*^ *or e*^*∗∗*^.

Proof of Proposition 1 can be found in the Appendix.

Once the hazard function is obtained, the following theorem shows how to obtain the different effects.

#### Theorem 1

*Under the conditions described in Proposition 1, it holds that the indirect effect of the mediator M*_*k*_, *k* = 1, …, *K is:*

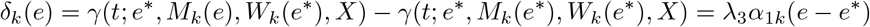

*Moreover, the joint indirect effect of all mediators Z is the sum of individual mediated effects:*

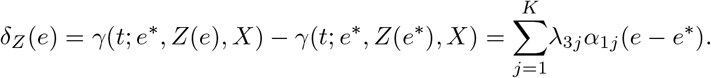

*The direct effect is:*

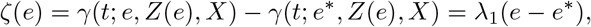

*and the total effect equals the sum of the joint indirect effect and the direct effect:*

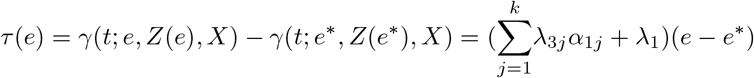

*Please note that, if we consider e*^*∗*^ = 0 *and e* = 1, *the factor* (*e − e*^*∗*^) *can be removed in all formulas. In addition, please note that δ*_*k*_(1) = *−δ*_*k*_(0), *δ*_*Z*_(1) = *−δ*_*Z*_(0), *ζ*(1) = *−ζ*(0) *and τ* (1) = *−τ* (0).

Proof of Theorem 1 is immediate as all effects correspond to differences in hazard functions, thus, the *C*(*t*) part is cancelled, and the other addends are also cancelled in the cases in which *e* = *e*^*∗*^, *e* = *e*^*∗∗*^ or *e*^*∗*^ = *e*^*∗∗*^. Please note that, given that *C*(*t*) is cancelled, all effects are independent of time.

## 4 Multimediate algorithm in survival settings

We used an adapted version of the quasi-bayesian algorithm developed in Jerolon et al. [7] to obtain point estimates of the effects of interest as well as confidence intervals and p-values. Let us consider the scenario of *K* mediators and *n* observations.

1. We fit the observed mediator model using linear regression, and the observed outcome model using the Lin-Ying model fitted with the *aalen* function from the R package *timereg*, which allows to specify that all coefficients are time-invariant except the baseline hazard.
2. We estimate the covariance matrix Σ of the errors of the mediator models by extracting the residuals 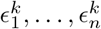 for each of the *K* mediator models and computing pairwise correlations between 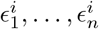 and 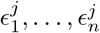 for each *i ≠ j*, obtaining the matrix 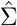. This matrix will be used later to incorporate the correlations between mediators to the simulation algorithm.
3. For each parameter of each of the models, we sample *J* values from the multivariate sampling distribution of their maximum likelihood estimators: 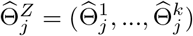 for the mediator models and 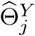 for the outcome model. For the mediator models, we use the multivariate normal distribution. For the Aalen model, the baseline hazard is not taken into account as all effect estimations imply a substraction in which the baseline hazard is cancelled (see section 3). According to Lin and Ying [22], all coefficients of the additive hazards model are also asymptotically normal. Thus, we also sample from the multivariate normal distribution for the outcome model. We use the estimates of the parameters as the mean, and the asymptotic covariance matrix between the estimators as the covariance.
4. In order to take into account the correlations between mediators, we jointly simulate the residuals of all the mediator models using a multivariate normal distribution with mean zero and covariance matrix 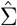.
5. For each simulation *j* = 1, …, *J*:
  a. We calculate the counterfactual values of each mediator under each exposure or treatment. For each of the *K* mediators, each pair of exposures (*e, e*^*∗*^) *∈ {*0, 1*}*^2^ and each individual *i* = 1, … *n*; 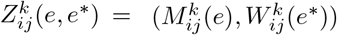.
  b. Given the simulated values of the counterfactual mediators, we calculate the counterfactual outcomes, i.e., for each individual *i*, mediator *k* and (*e, e*^*∗*^, *e*^*∗∗*^) *∈ {*0, 1*}*^3^, we calculate 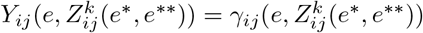.
  c. We estimate the causal mediation effects. Our proposed estimators are the sample mean of the effects obtained in the previous simulation process:
    - Indirect effect for each mediator:

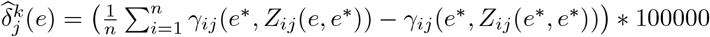
    - Joint indirect effect:

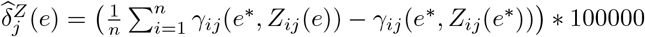
    - Direct effect:

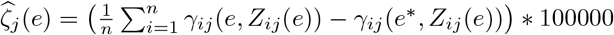
    - Total effect:

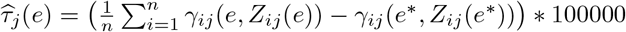

Each effect is calculated for both *e* = 0 and *e* = 1. In section 3, we proved that *δ*_*k*_(1) = *−δ*_*k*_(0) but, in general, 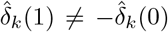, as they represent two different estimators of the same parameter. Thus, we propose to use 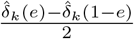 as the estimator of 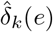. Similarly for direct and total effects. Please note that we multiply each estimator by 100, 000 in order to get an estimation of the number of cases attributable to the exposure through the mediator per 100, 000 person-years (this number could be changed according to the users preferences). Also note that, for time-invariant covariates, the effects do not depend on the time *t*.
6. From the empirical distribution of each effect above, we obtain confidence intervals: The 50-th percentile is taken as the average effect of interest, and the 2.5-th and 97.5-th percentiles of the sample distribution of each estimator are taken as the 95 % confidence intervals’ lower and upper bounds, respectively.

## 5 A simulation study

We conducted a simulation study in order to assess the performance of the multimediate algorithm in survival settings, and compare it to simple mediation analysis. For the purposes of this simulation study, we assume the setting of three mediators (*K* = 3). Following Jerolon et al.’s [7] simulation framework, we first simulate a database of 10^6^ observations for exposure *e ∈ {*0, 1*}*, for the counterfactual mediators *M*_1_, *M*_2_ and *M*_3_ and the counterfactual value of the linear predictor 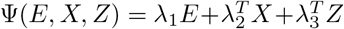 (hereinafter referred to as Ψ for simplicity), which equals the definition of the rate *γ* in additive models except for the baseline hazard, which is removed as all effect calculations require substractions and the baseline hazard is cancelled. We will subsequently use this linear predictor to calculate survival times for each individual. We then calculate the direct, indirect and total effects as described in section 3, substracting means of the counterfactual values of the linear predictor in different scenarios. The large size of the database guarantees that those estimates are sufficiently close to the true values of the effects. We fixed the number of simulations to 600. In each simulation, a random sample of 2000 observations of the full database is taken, and the effects of interest are calculated in that subsample.

The Mean Squared Error (MSE), the bias, the variance and the % coverage of the 95 % confidence intervals are calculated comparing the true effects (calculated in the full simulated database) to those estimated by simple mediation analysis and by the multimediate algorithm.

In order to simulate survival times, we use the inverse transformation method. Please note that the survival distribution function is *S*(*t*) = *exp*(*−*Λ(*t*)), being Λ(*t*) the cumulative hazard function, in our case 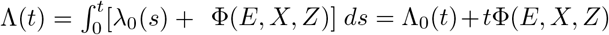, where Λ_0_(*t*) is the cumulative baseline hazard function. Hence, a simulated time *t* is obtained as the solution of the equation *u* = *exp*(*−*Λ_0_(*t*) *− t*Φ(*E, X, Z*)), being *u* a number randomly generated from a *U* (0, 1) distribution [23].

We consider three different scenarios for the baseline hazard: constant baseline hazard, monotonic baseline hazard dependent on time, and non-monotonic baseline hazard. In addition, we consider three different correlation scenarios for the mediator: negative correlation (*ρ* = *−*0.4), no correlation (*ρ* = 0) and positive correlation (*ρ* = 0.4). In the next sections, we present the results of the metrics (MSE, bias, variance and confidence interval coverage) of the simulations in each of the scenarios.

### 5.1 Constant baseline hazard

We assume that the baseline hazard takes the constant value *λ*_0_ = 0.1. Given that in this case Λ_0_(*t*) = 0.1*t*, the survival time for a given individual would be simulated as:

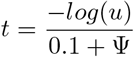

being *u ∼ U* (0, 1). Tables 1, 2 and 3 show the Mean Squared Errors (MSE), variance and bias for the total, direct and indirect effects comparing simple mediation to the multimediate algorithm. While both frameworks present similar results for the total effect, the multimediate algorithm presents, in general, a smaller MSE for the direct effect, even in the setting of no correlation between mediators. For the indirect effect, although the error is similar in the context of no correlation between mediators, the error is smaller for the multimediate algorithm in contexts of both positive and negative correlations between mediators. The reduction in bias of the multimediate algorithm drives the reduction in MSE.

**Table 1:** Simulation results for total effect in a constant baseline risk scenario.

|  | Correlation = -0.4 |  |  |  | Correlation = 0 |  |  |  | Correlation = 0.4 |  |  |  |
| --- | --- | --- | --- | --- | --- | --- | --- | --- | --- | --- | --- | --- |
|  | Med 1 | Med 2 | Med 3 | Multim | Med 1 | Med 2 | Med 3 | Multim | Med 1 | Med 2 | Med 3 | Multim |
| MSE | 0.23 | 0.23 | 0.23 | 0.23 | 0.26 | 0.26 | 0.26 | 0.27 | 0.25 | 0.25 | 0.25 | 0.25 |
| Var | 0.23 | 0.23 | 0.23 | 0.23 | 0.26 | 0.26 | 0.26 | 0.27 | 0.25 | 0.25 | 0.25 | 0.25 |
| Bias | -0.033 | -0.034 | -0.035 | -0.033 | -0.029 | -0.029 | -0.027 | -0.029 | -0.012 | -0.011 | -0.013 | -0.014 |

**Table 2:** Simulation results for direct effect in a constant baseline risk scenario.

|  | Correlation = -0.4 |  |  |  | Correlation = 0 |  |  |  | Correlation = 0.4 |  |  |  |
| --- | --- | --- | --- | --- | --- | --- | --- | --- | --- | --- | --- | --- |
|  | Med 1 | Med 2 | Med 3 | Multim | Med 1 | Med 2 | Med 3 | Multim | Med 1 | Med 2 | Med 3 | Multim |
| MSE | 0.68 | 1.79 | 0.72 | 0.71 | 0.51 | 0.95 | 0.50 | 0.36 | 0.39 | 0.47 | 0.35 | 0.31 |
| Var | 0.26 | 0.30 | 0.25 | 0.71 | 0.29 | 0.29 | 0.29 | 0.36 | 0.28 | 0.29 | 0.28 | 0.31 |
| Bias | 0.65 | 1.22 | 0.68 | -0.014 | 0.47 | 0.81 | 0.46 | -0.048 | 0.33 | 0.42 | 0.26 | -0.005 |

**Table 3:** Simulation results for indirect effects (simple mediation / multimediate) in a constant baseline risk scenario.

|  | Correlation = -0.4 |  |  |
| --- | --- | --- | --- |
|  | Med 1 | Med 2 | Med 3 |
| MSE | 0.056 / 0.049 | 0.21 / 0.074 | 0.078 / 0.063 |
| Var | 0.022 / 0.049 | 0.039 / 0.075 | 0.031 / 0.063 |
| Bias | -0.18 / -0.019 | -0.42 / -0.0027 | -0.22 / 0.002 |
|  | Correlation = 0 |  |  |
|  | Med 1 | Med 2 | Med 3 |
| MSE | 0.026 / 0.026 | 0.038 / 0.037 | 0.029 / 0.030 |
| Var | 0.026 / 0.026 | 0.038 / 0.038 | 0.029 / 0.030 |
| Bias | -0.0007 / -0.003 | -0.0015 / 0.0031 | 0.016 / 0.019 |
|  | Correlation = 0.4 |  |  |
|  | Med 1 | Med 2 | Med 3 |
| MSE | 0.051 / 0.031 | 0.21 / 0.051 | 0.084 / 0.039 |
| Var | 0.025 / 0.031 | 0.042 / 0.051 | 0.034 / 0.039 |
| Bias | 0.16 / -0.011 | 0.40 / -0.0068 | 0.22 / 0.0094 |

Tables 4 and 5 show the empirical coverage of 95 % confidence intervals in terms of proportions of simulations that contain the real value of the different effects (calculated in the full database of 1,000,000 observations). While the total effect has great empirical coverage for both simple mediation and the multimediate algorithm, direct and indirect effects clearly worsen their empirical coverage in simple mediation models in settings of correlated mediators. Conversely, the multimediate algorithm remains with good and similar coverage in both correlated and uncorrelated settings.

**Table 4:** Empirical coverage of the confidence interval with theoretical coverage of 95 % (in proportions of simulations) of simple mediation models in a constant baseline risk scenario.

|  | Mediator 1 |  |  |
| --- | --- | --- | --- |
|  | Indirect | Direct | Total |
| Correlation=-0.4 | 0.76 | 0.75 | 0.96 |
| Correlation=0 | 0.95 | 0.84 | 0.95 |
| Correlation=0.4 | 0.82 | 0.90 | 0.95 |
|  | Mediator 2 |  |  |
|  | Indirect | Direct | Total |
| Correlation=-0.4 | 0.44 | 0.37 | 0.96 |
| Correlation=0 | 0.97 | 0.67 | 0.95 |
| Correlation=0.4 | 0.46 | 0.87 | 0.95 |
|  | Mediator 3 |  |  |
|  | Indirect | Direct | Total |
| Correlation=-0.4 | 0.76 | 0.75 | 0.96 |
| Correlation=0 | 0.95 | 0.85 | 0.95 |
| Correlation=0.4 | 0.75 | 0.92 | 0.95 |

**Table 5:** Empirical coverage of the confidence interval with theoretical coverage of 95 % (in proportions of simulations) of the multimediate algorithm in a constant baseline risk scenario.

|  | Indirect M1 | Indirect M2 | Indirect M3 | Direct | Total |
| --- | --- | --- | --- | --- | --- |
| Correlation=-0.4 | 0.94 | 0.96 | 0.94 | 0.95 | 0.94 |
| Correlation=0 | 0.94 | 0.95 | 0.95 | 0.93 | 0.93 |
| Correlation=0.4 | 0.93 | 0.92 | 0.95 | 0.94 | 0.95 |

### 5.2 Monotonic baseline hazard dependent on time

We now assume that the baseline hazard takes the value *λ*_0_ = *t*. Thus, the cumulative hazard function would be defined as:

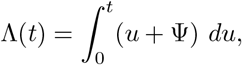

and the survival function would be defined as:

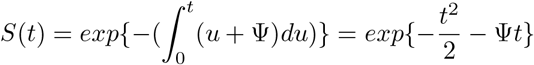

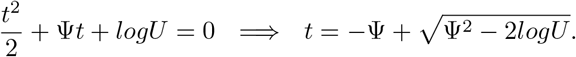

Please note that, given that 0 *< U <* 1, it always holds that 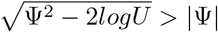. Therefore, 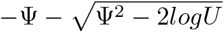 is not considered as a possible solution as survival times are always positive.

Tables 6, 7 and 8 show the Mean Squared Errors (MSE), variance and bias for the total, direct and indirect effects comparing simple mediation to the multimediate algorithm. A similar tendency to that of the constant baseline hazard case can be observed. Again, both frameworks present similar results for the total effect and the multimediate algorithm presents, in general, a smaller MSE for the direct effect, even in the context of no correlation between mediators. For the indirect effect, the error is again similar in the context of no correlation between mediators, and smaller for the multimediate algorithm in contexts of correlated mediators.

**Table 6:** Simulation results for total effect in a monotonic time-dependent baseline risk scenario.

|  | Correlation = -0.4 |  |  |  | Correlation = 0 |  |  |  | Correlation = 0.4 |  |  |  |
| --- | --- | --- | --- | --- | --- | --- | --- | --- | --- | --- | --- | --- |
|  | Med 1 | Med 2 | Med 3 | Multim | Med 1 | Med 2 | Med 3 | Multim | Med 1 | Med 2 | Med 3 | Multim |
| MSE | 64.8 | 64.9 | 64.5 | 64.1 | 62.1 | 62.5 | 62.5 | 61.4 | 66.3 | 66.5 | 66.3 | 65.4 |
| Var | 50.8 | 50.9 | 50.6 | 50.9 | 48.1 | 48.1 | 48.1 | 48.1 | 50.1 | 50.4 | 50.4 | 49.9 |
| Bias | -3.7 | -3.7 | -3.7 | -3.6 | -3.7 | -3.8 | -3.8 | -3.7 | -4.0 | -4.0 | -4.0 | -3.9 |

**Table 7:** Simulation results for direct effect in a monotonic time-dependent baseline risk scenario.

|  | Correlation = -0.4 |  |  |  | Correlation = 0 |  |  |  | Correlation = 0.4 |  |  |  |
| --- | --- | --- | --- | --- | --- | --- | --- | --- | --- | --- | --- | --- |
|  | Med 1 | Med 2 | Med 3 | Multim | Med 1 | Med 2 | Med 3 | Multim | Med 1 | Med 2 | Med 3 | Multim |
| MSE | 4551.9 | 15205.0 | 5191.4 | 5882.7 | 2587.5 | 6769.4 | 2610.5 | 1456.6 | 1455.2 | 2349.3 | 1168.5 | 879.3 |
| Var | 360.0 | 671.3 | 499.3 | 5887.6 | 364.9 | 626.1 | 537.7 | 1425.0 | 375.9 | 661.3 | 515.3 | 880.7 |
| Bias | 64.7 | 120.6 | 68.5 | -2.2 | 47.1 | 78.4 | 45.5 | -5.8 | 32.9 | 41.1 | 25.6 | 0.30 |

**Table 8:** Simulation results for indirect effects (simple mediation / multimediate) in a monotonic time-dependent baseline risk scenario.

|  | Correlation = -0.4 |  |  |
| --- | --- | --- | --- |
|  | Med 1 | Med 2 | Med 3 |
| MSE | 650.4 / 641.3 | 2236.9 / 1109.2 | 949.6 / 872.9 |
| Var | 308.8 / 639.0 | 613.1 / 1110.8 | 455.5 / 874.3 |
| Bias | -18.5 / -1.8 | -40.3 / 0.56 | -22.2 / -0.16 |
|  | Correlation = 0 |  |  |
|  | Med 1 | Med 2 | Med 3 |
| MSE | 309.2 / 310.3 | 598.4 / 597.8 | 491.0 / 492.6 |
| Var | 308.9 / 309.7 | 596.1 / 594.3 | 491.4 / 492.3 |
| Bias | -0.91 / -1.1 | 1.8 / 2.1 | 0.67 / 1.09 |
|  | Correlation = 0.4 |  |  |
|  | Med 1 | Med 2 | Med 3 |
| MSE | 508.3 / 458.6 | 2114.0 / 728.9 | 874.0 / 575.9 |
| Var | 337.1 / 445.5 | 603.5 / 729.5 | 457.7 / 576.9 |
| Bias | 13.1 / -3.7 | 38.9 / -0.76 | 20.4 / 0.26 |

Tables 9 and 10 show the empirical coverage of 95 % confidence intervals. As for the constant baseline risk scenario, total effects have similar empirical coverage for both simple mediation and the multimediate algorithm. However, the empirical coverage is much better for the multimediate algorithm for both direct and indirect effects. Direct effects have sometimes null empirical coverage in the simple mediation models, and the empirical coverage is also clearly worse in contexts of correlated settings. The multimediate model maintains good and similar empirical coverage for all effects.

**Table 9:** Empirical coverage of the confidence interval with theoretical coverage of 95 % (in proportions of simulations) of simple mediation models in a monotonic time-dependent baseline risk scenario.

|  | Mediator 1 |  |  |
| --- | --- | --- | --- |
|  | Indirect | Direct | Total |
| Correlation=-0.4 | 0.81 | 0.07 | 0.91 |
| Correlation=0 | 0.95 | 0.32 | 0.92 |
| Correlation=0.4 | 0.88 | 0.59 | 0.89 |
|  | Mediator 2 |  |  |
|  | Indirect | Direct | Total |
| Correlation=-0.4 | 0.60 | 0.005 | 0.91 |
| Correlation=0 | 0.94 | 0.12 | 0.91 |
| Correlation=0.4 | 0.60 | 0.60 | 0.90 |
|  | Mediator 3 |  |  |
|  | Indirect | Direct | Total |
| Correlation=-0.4 | 0.81 | 0.11 | 0.91 |
| Correlation=0 | 0.94 | 0.46 | 0.92 |
| Correlation=0.4 | 0.81 | 0.76 | 0.91 |

**Table 10:** Empirical coverage of the confidence interval with theoretical coverage of 95 % (in proportions of simulations) of the multimediate algorithm in a monotonic time-dependent baseline risk scenario.

|  | Indirect M1 | Indirect M2 | Indirect M3 | Direct | Total |
| --- | --- | --- | --- | --- | --- |
| Correlation=-0.4 | 0.96 | 0.94 | 0.94 | 0.95 | 0.90 |
| Correlation=0 | 0.95 | 0.92 | 0.93 | 0.94 | 0.90 |
| Correlation=0.4 | 0.92 | 0.94 | 0.95 | 0.93 | 0.89 |

### 5.3 Non-monotonic baseline hazard

Let us now define the baseline hazard as the following piecewise function:

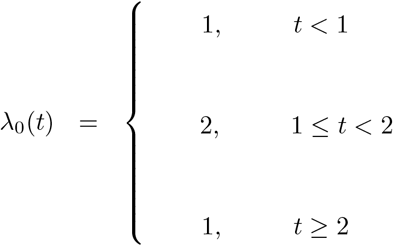

Then, the cumulative risk would be defined as:

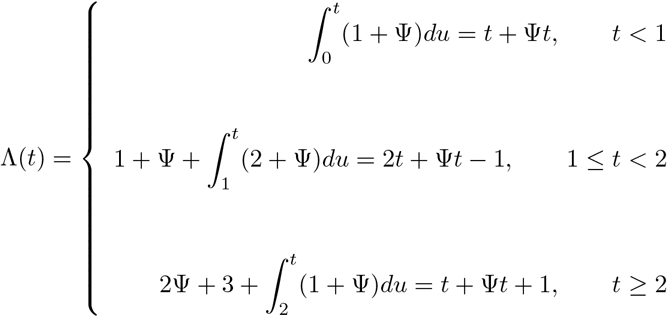

Thus, the survival function would be defined as:

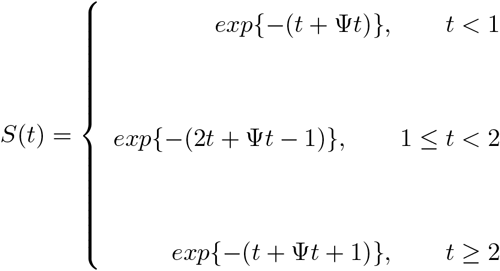

and, following simple inequalities calculations, the survival time t would be simulated as:

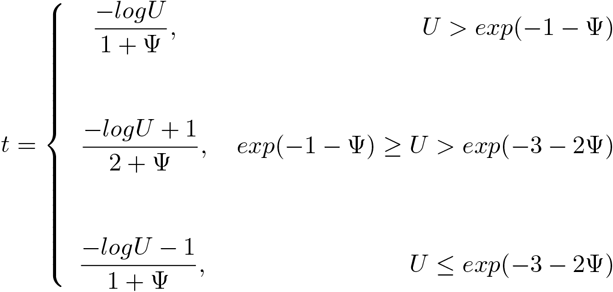

Tables 11, 12 and 13 show the Mean Squared Errors (MSE), variance and bias for the total, direct and indirect effects comparing simple mediation to the multimediate algorithm. Tables 14 and 15 show the empirical coverage of confidence intervals. The patterns are essentially similar to those observed in the previous two baseline hazard scenarios.

**Table 11:** Simulation results for total effect in a non-monotonic baseline risk scenario.

|  | Correlation = -0.4 |  |  |  | Correlation = 0 |  |  |  | Correlation = 0.4 |  |  |  |
| --- | --- | --- | --- | --- | --- | --- | --- | --- | --- | --- | --- | --- |
|  | Med 1 | Med 2 | Med 3 | Multim | Med 1 | Med 2 | Med 3 | Multim | Med 1 | Med 2 | Med 3 | Multim |
| MSE | 44.8 | 45.1 | 44.9 | 45.1 | 43.5 | 43.5 | 43.4 | 43.6 | 40.9 | 40.7 | 40.7 | 40.9 |
| Var | 44.9 | 45.1 | 44.9 | 45.0 | 43.6 | 43.5 | 43.4 | 43.6 | 40.9 | 40.8 | 40.8 | 40.9 |
| Bias | 0.27 | 0.28 | 0.28 | 0.34 | 0.11 | 0.11 | 0.10 | 0.18 | 0.06 | 0.11 | 0.07 | 0.17 |

**Table 12:** Simulation results for direct effect in a non-monotonic baseline risk scenario.

|  | Correlation = -0.4 |  |  |  | Correlation = 0 |  |  |  | Correlation = 0.4 |  |  |  |
| --- | --- | --- | --- | --- | --- | --- | --- | --- | --- | --- | --- | --- |
|  | Med 1 | Med 2 | Med 3 | Multim | Med 1 | Med 2 | Med 3 | Multim | Med 1 | Med 2 | Med 3 | Multim |
| MSE | 3620.3 | 1603.6 | 3066.6 | 2135.9 | 2264.4 | 669.3 | 2639.5 | 605.5 | 1298.5 | 584.7 | 2329.7 | 586.6 |
| Var | 122.3 | 505.2 | 55.3 | 2098.3 | 124.9 | 498.2 | 50.1 | 606.3 | 126.4 | 539.0 | 52.5 | 586.7 |
| Bias | 59.1 | 33.2 | 54.9 | 6.4 | 46.3 | 13.1 | 50.9 | -0.43 | 34.2 | -6.8 | 47.7 | -0.94 |

**Table 13:** Simulation results for indirect effects (simple mediation / multimediate) in a non-monotonic baseline risk scenario.

|  | Correlation = -0.4 |  |  |
| --- | --- | --- | --- |
|  | Med 1 | Med 2 | Med 3 |
| MSE | 253.8 / 201.6 | 846.5 / 988.8 | 21.8 / 20.9 |
| Var | 88.3 / 196.9 | 452.2 / 979.1 | 8.8 / 20.7 |
| Bias | -12.9 / -2.3 | -19.9 / -3.4 | -3.6 / -0.46 |
|  | Correlation = 0 |  |  |
|  | Med 1 | Med 2 | Med 3 |
| MSE | 88.5 / 89.1 | 445.0 / 450.4 | 8.5 / 8.5 |
| Var | 88.6 / 89.2 | 445.7 / 450.9 | 8.4 / 8.4 |
| Bias | -0.15 / -0.23 | 0.001 / 0.57 | 0.21 / 0.27 |
|  | Correlation = 0.4 |  |  |
|  | Med 1 | Med 2 | Med 3 |
| MSE | 224.1 / 105.2 | 874.6 / 617.9 | 20.9 / 12.5 |
| Var | 84.5 / 105.4 | 478.0 / 616.6 | 9.7 / 12.4 |
| Bias | 11.8 / -0.11 | 19.9 / 1.5 | 3.3 / -0.3 |

**Table 14:** Empirical coverage of the confidence interval with theoretical coverage of 95 % (in proportions of simulations) of simple mediation models in a non-monotonic baseline risk scenario.

|  | Mediator 1 |  |  |
| --- | --- | --- | --- |
|  | Indirect | Direct | Total |
| Correlation=-0.4 | 0.71 | 0 | 0.96 |
| Correlation=0 | 0.94 | 0.01 | 0.95 |
| Correlation=0.4 | 0.73 | 0.15 | 0.96 |
|  | Mediator 2 |  |  |
|  | Indirect | Direct | Total |
| Correlation=-0.4 | 0.83 | 0.66 | 0.96 |
| Correlation=0 | 0.96 | 0.91 | 0.95 |
| Correlation=0.4 | 0.83 | 0.92 | 0.96 |
|  | Mediator 3 |  |  |
|  | Indirect | Direct | Total |
| Correlation=-0.4 | 0.78 | 0 | 0.96 |
| Correlation=0 | 0.97 | 0 | 0.95 |
| Correlation=0.4 | 0.81 | 0 | 0.96 |

**Table 15:** Empirical coverage of the confidence interval with theoretical coverage of 95 % (in proportions of simulations) of the multimediate algorithm in a non-monotonic baseline risk scenario.

|  | Indirect M1 | Indirect M2 | Indirect M3 | Direct | Total |
| --- | --- | --- | --- | --- | --- |
| Correlation=-0.4 | 0.93 | 0.94 | 0.94 | 0.94 | 0.95 |
| Correlation=0 | 0.94 | 0.93 | 0.97 | 0.94 | 0.94 |
| Correlation=0.4 | 0.95 | 0.93 | 0.94 | 0.93 | 0.95 |

## 6 Discussion

In this work, we extended the quasi-bayesian multimediate algorithm to a time-to-event setting using the semiparametric additive hazards model. We theoretically demonstrated that, under certain assumptions, indirect, direct and total effects can be calculated using the counterfactual framework with multiple correlated mediators. We additionally conducted a simulation study under different baseline risk scenarios and different levels of correlations between mediators to show that the multimediate algorithm has a better performance than simple mediation analysis using the product of coefficients method, especially in the setting in which mediators are correlated. This work has been added to the Github repository *https://github.com/AllanJe/multimediate* as part of an extension of the original R package *multimediate* developed by [7].

Our simulation study shows that, in general, and regardless of the baseline risk definition, the mean squared errors are smaller for both direct and indirect effects for the multimediate algorithm as compared to those of the simple mediation framework, especially in settings of correlated mediators. Of note, the empirical coverage of the confidence intervals in the multimediate algorithm is far better than that of simple mediation analysis, in which the empirical coverage is worsened for both direct and indirect effects in the context of correlated mediators.

Survival analysis is widely used in mediation analysis applied to medical settings, in which one might be interested in evaluating the potential mediating effect of a biological process on the association between an exposure or treatment and a health outcome. Incorporating information on the time in which individuals developed a health event is essential to accurately evaluate disease risk. Traditionally, mediation analysis has been conducted using Aalen additive hazards models [13], however, to our knowledge, no multi-mediator algorithms for correlated mediators with survival endpoints have been developed to date. Aalen additive hazards models have several advantages as compared to Cox proportional hazards models. Rate differences provide a more straightforward interpretation in attributable cases per person-years and, unlike hazard ratios, are collapsible [19], meaning that the magnitude of the coefficient of the exposure would not change when adjusting the model for a variable that is unrelated to the exposure. In addition, in settings in which the proportional hazards assumption is not fulfilled [24], the Aalen model is more appropriate.

However, this model is not without complications. Convergency issues might arise with this survival version of the multimediate algorithm in settings of small sample sizes or very high inverse correlations between mediators, as the Aalen model might present more convergency issues than the Cox model. In our setting, inverse correlations between mediators lower than *−*0.4 presented convergency issues even for sample sizes greater than 10, 000. On the other hand, it is known that, given that the Aalen model and survival models in general are less informative than linear models due to censoring, larger sample sizes are needed for a survival model than for a linear model to obtain similar results in terms of robustness. This is the reason why we chose larger sample sizes for the simulation study as compared to the simulation study conducted in Jerolon et al. for continuous outcomes [7]. Future work would need to extend this algorithm to accelerated failure time models in order to be able to choose the adequate survival analysis method depending on the framework.

Furthermore, the context of this work requires two important assumptions. First, as stated in Jerolon et al. [7], this work is restricted to the setting in which the correlation between counterfactual mediators is independent of the exposure or treatment. Relevant future work should include the development of methods for addressing the situation in which the correlation between mediators is dependent on the exposure. Second, we assume that the joint distribution of the mediators is a multivariate normal. This is not necessarily true in settings in which mediators are not independent. However, this is not feasible to prove in practice as all linear combinations of the mediators should follow a normal distribution in order to conclude that the joint distribution of the mediators is a multivariate normal. Deviations from multivariate normality should be studied in future work.

Of note, the multimediate algorithm uses the counterfactual framework to identify direct, indirect and total effects. Traditional mediation approaches such as the product of coefficients and the difference of coefficients [20] approaches can lead to biased effect estimates in presence of exposure-mediator interactions. As stated by [9], the natural direct effects and natural indirect effects as defined by the counterfactual framework can provide valid estimates even in the case of exposure-mediator interactions. Our extension of the *multimediate* algorithm provides direct, indirect and total effect estimates in all strata of the exposure, thus, potential exposure-mediator interactions can be identified.

Importantly, the no unmeasured confounding assumptions in the exposure-mediator, exposure-outcome and mediator-outcome relationships reminds a fundamental issue to be able to identify valid effects in mediation analysis. Several sensitivity analyses to identify and even correct for measurement errors and unmeasured confounding have been developed [4, 17, 25, 16]. Developing sensitivity analyses that illustrate the potential effect that hypothetical unmeasured confounders would need to have in order to explain the whole direct, indirect or total effects in the multimediate algorithm should also constitute relevant future work.

In conclusion, the multimediate algorithm is able to conduct multiple mediation in presence of correlations between mediators. Unlike multiplicative models, the semiparametric additive risks model provides the effect in a rate difference scale, which is a more interpretable measure in a survival setting and can be highly informative for public health.

## Data Availability

All data produced in the present study were simulated.

## Acknowledgments

ADR was supported by a fellowship from “la Caixa” Foundation (ID 100010434) (fellowship code “LCF/BQ/DR19/11740016”), and by the National Institute of Environmental Health Sciences of the United States (P42ES033719). Dr. Tellez-Plaza received funding from the Strategic Action for Research in Health sciences (CP12/03080 and PI15/00071), which are initiatives from Instituto de Salud Carlos III and the Spanish Ministry of Science and Innovation and co-funded with European Funds for Regional Development (FEDER) and by the State Agency for Research (PID2019-108973RB-C21).

