## Appendix for "Causal mediation for uncausally related mediators in the context of survival analysis"

### Appendix: Proof of Proposition 1

Without loss of generality, we consider $k = 1$. Let us call $T^*$ to the random variable $T(e, M_1(e^*), W_1(e^{**}))$, being $(e, e^*, e^{**}) \in \{0, 1\}^3$. Then, the rate can be expressed as:

$$\gamma(t; e, M_1(e^*), W_1(e^{**})) = \lim_{dt \to 0} \frac{1}{dt}\ P(T^* \in [t, t + dt] \mid T^* \geq t).$$

It holds that:

$$P(T^* \in [t, t + dt] \mid T^* \geq t) = \mathbb{E}_{\{X|T^*>t\}}[P(T^* \in [t, t + dt] \mid X = x, T^* \geq t)]$$

and, similarly, being $F(m_1, w_1)$ the distribution function of $M_1(e^*), W_1(e^{**})$ given that $X = x$ and $T^* > t$:

$$\begin{aligned} P(T^* \in [t, t + dt]|X = x, T^* \geq t) &= \textstyle\int_{\mathbb{R}^K} P(T^* \in [t, t + dt] \mid X = x, M_1 = m_1, W_1 = w_1, T^* \geq t)\ dF(m_1, w_1) \\ &= \textstyle\int_{\mathbb{R}^K} P(T(e, m_1, w_1) \in [t, t + dt] \mid X = x, M_1 = m_1, W_1 = w_1, T(e, m_1, w_1) > t)\ dF(m_1, w_1) \\ &= \textstyle\int_{\mathbb{R}^K} P(T(e, m_1, w_1) \in [t, t + dt]|X = x, E = e, M_1 = m_1, W_1 = w_1, T(e, m_1, w_1) > t) dF(m_1, w_1). \end{aligned}$$

Using the bounded convergency theorem and the additive risk hypothesis:

$$\begin{aligned} \gamma(t; e, M_1(e^*), W_1(e^{**})) &= \mathbb{E}_{\{X|T^*>t\}}\Big[\int_{\mathbb{R}^k} (\lambda_0(t) + \lambda_1 e + \lambda_2^T x + \lambda_3^T (m_1, w_1^T)^T)\ dF(m_1, w_1)\Big] \\ &= \lambda_o(t) + \lambda_1 e + \lambda_2^T\ \mathbb{E}(X|T^* > t) + \mathbb{E}_{\{X|T^*>t\}}\Big[\int_{\mathbb{R}^K} \lambda_3^T (m_1, w_1^T)^T\ dF(m_1, w_1)\Big]. \end{aligned}$$

In addition,

$$\begin{aligned} \textstyle\int_{\mathbb{R}^K} \lambda_3^T (m_1, w_1^T)^T dF(m_1, w_1) &= \textstyle\int_{\mathbb{R}^K} \lambda_3^T (m_1, w_1^T)^T\ f(M_1(e^*) = m_1, W_1(e^{**}) = w_1 \mid X = x, T^* > t)\ dm_1 dw_1 \\ &= \textstyle\int_{\mathbb{R}^K} \lambda_3^T (m_1, w_1^T)^T\ \frac{P(T^*>t \mid M_1=m_1, W_1=w_1, X=x)\ f(M_1(e^*)=m_1, W_1(e^{**})=w_1 \mid X=x)}{P(T^*>t|X=x)}\ dm_1 dw_1. \end{aligned}$$

Following the same arguments and taking into account that Aalen additive hazards models have been used for a time-to-event setting:

$$\begin{aligned} P(T^* > t \mid M_1 = m_1, W_1 = w_1, X = x) &= P(T(e, m_1, w_1) > t \mid M_1 = m_1, W_1 = w_1, X = x, E = e) \\ &= exp\{-\int_0^t \lambda_0(u)du - \lambda_1 et - \lambda_2^T xt - \lambda_3^T (m_1, w_1^T)^T t\}. \end{aligned}$$

Also,

$$\begin{aligned} P(T^* > t|X = x) &= \int_{\mathbb{R}^K} P(T^* > t \mid M_1 = m_1, W_1 = w_1, X = x)\ dF(m_1, w_1) \\ &= exp\Big\{-\int_0^t \lambda_0(u)du - \lambda_1 et - \lambda_2^T xt\Big\}\ \mathbb{E}\ (exp\{-\lambda_3^T (m_1, w_1^T)^T t\}) \end{aligned}$$

Hence, defining $V = \lambda_3^T (m_1, w_1^T)^T$ and putting together the above results, it holds that:

$$\int_{\mathbb{R}^K} \lambda_3^T (m_1, w_1^T)^T\ dF(m_1, w_1) = \int_{\mathbb{R}^K} V\ dF(m_1, w_1) = \frac{\mathbb{E}(V exp\{-tV\} \mid X = x)}{\mathbb{E}(exp\{-tV\} \mid X = x)}.$$

The distribution of $(M_1(e^*), W_1(e^{**}) \mid X = x)$ is multivariate normal [**?**] with covariance matrix $\Sigma$, which does not depend on the exposures, and expected values:

$\mathbb{E}(M_1(e^*) \mid X = x) = \alpha_{01} + \alpha_{11} e^* + \alpha_{21} x$

$\mathbb{E}(M_j(e^{**}) \mid X = x) = \alpha_{0j} + \alpha_{1j} e^{**} + \alpha_{2j} x,\ j = 2, ..., K.$

Thus, the distribution of $V$ is normal with:

$\mathbb{E}(V \mid X = x) = \lambda_3^T \alpha_0 + \lambda_{31}\alpha_{11} e^* + \sum_{j=2}^{K} \lambda_{3j}\alpha_{1j} e^{**} + \lambda_3^T \alpha_2 x$

$Var(V \mid X = x) = \lambda_3^T \Sigma \lambda_3.$

Thus, following Lange and Hansen 2011 [**?**],

$$\begin{aligned}\frac{\mathbb{E}(Vexp\{-tV\}|X=x)}{\mathbb{E}(exp\{-tV\}|X=x)} &= \mathbb{E}(V|X=x) - tVar(V|X=x) \\ &= \lambda_3^T\alpha_0 + \lambda_{31}\alpha_{11}e^* + \sum_{j=2}^{K}\lambda_{3j}\alpha_{1j}e^{**} + \lambda_3^T\alpha_2 x - t\lambda_3^T\Sigma\lambda_3,\end{aligned}$$

and it holds that the counterfactual rate can be expressed as:

$$\gamma(t; e, M(e^*), W(e^{**})) = C(t) + \lambda_1 e + \lambda_{31}\alpha_{11}e^* + \sum_{j=2}^{K}\lambda_{3j}\alpha_{1j}e^{**},$$

being $C(t) = \lambda_0(t) + \lambda_2^T\ \mathbb{E}(X \mid T^* > t) + \lambda_3^T\alpha_0 + \lambda_3^T\alpha_2\ \mathbb{E}(X \mid T^* > t) - t\lambda_3^T\Sigma\lambda_3$ a function of $t$ that does not depend on the exposures.
